# Morphological alternations of intracerebral arterial during middle age: a community cohort study

**DOI:** 10.1101/2021.09.13.21263492

**Authors:** Boyu Zhang, Zidong Yang, Jing Li, Bei Wang, Huazheng Shi, He Wang, Yuehua Li

## Abstract

**Background:** Ample evidence has suggested that vascular modifications are associated with aging. To expand previous understanding of age-related vascular changes, we examined the association between aging and cerebrovascular morphologies.

**Methods:** 1176 participants aged 35 to 75 years recruited from Shanghai, China were included in this study. Cerebrovascular morphological features comprising arterial branch density, radius, and tortuosity were quantified using three-dimensional magnetic resonance angiography. Linear regression was used to examine the association between age and vasculature features.

**Results:** Age was found to be a significant predictor for cerebrovascular morphological alterations after adjusting for vascular risk factors. However, after dividing subjects into subgroups based on their age, aging was found to be significantly correlated with all three morphometric features only in the 45-54 subgroup after adjusting for the other vascular risk factors. Smoking gives rise to a more rapid age-related changes in vascular morphologies, while alcohol consumption could decelerate those age-related alterations.

**Conclusions:** Rapid alternations in all three morphological features assessed have been noticed to be associated with aging in the 45-54 subgroup, suggesting the potential importance of the 5th decade for early preservation method of vascular aging.

**Funding:** National Natural Science Foundation of China.

## Introduction

The power-hungry nature of the human brain requires a consistent supply of oxygen and nutrients, which is dependent on the structural and functional integrity of the intracranial vasculature, to maintain its normal functioning (Zhang and Raichle 2010). Indeed, the cerebral vasculature has been implicated in many dementing diseases, such as vascular dementia and Alzheimer’s disease (AD) (Leeuwis et al. 2017, Alber et al. 2019). Several previous studies have suggested a synergistic relationship between cerebrovascular dysfunctions and the neurological symptoms (Silvestrini et al. 2006, Kalaria 2002). For instance, reduced cerebral blood flow (CBF) has been implicated in cognitive decline (Swan et al. 1998, Leeuwis et al. 2017). Exposure to cerebrovascular risk (CVR) could cause cerebral vascular system impairments. In particular, elevated blood pressure is associated with the remodeling of both small and large arteries, which paves the way for the impairment of the brain functions (Laurent and Boutouyrie 2015). Among the common CVR factors, aging has been recognized to pose a significant risk for the development of cardiovascular diseases, and its effect on the cerebral vascular system deserves more attention (D’Agostino et al. 2008).

Association between advanced age and a general reduction in CBF in normal subjects has been reported earlier (Melamed et al. 1980). During the past few years, clinical studies have revealed that major vascular modifications, including generalized endothelial dysfunction and central arterial stiffness, in the cerebral arteriolar system also transpire with aging (Xu et al. 2017, Donato, Machin and Lesniewski 2018). With the advancements in time-of-flight magnetic resonance angiography (TOF MRA), age-related intracranial arterial morphological alterations, including reduced vessel branch number and increased mean tortuosity have been reported(Chen et al. 2019, Bullitt et al. 2010). Such alterations have recently been implicated in cerebral small vessel disease (CSVD), a brain disease that is believed to be associated with a wide range of cognitive disorders including nearly a half of dementias, making those promising markers for brain damage and cognitive decline (Banerjee et al. 2016, Zhang et al. 2021).

Moreover, extensive research has focused on midlife since exposure to CVR factors during midlife was reported to influence cognitive outcomes later on significantly (Walker et al. 2019, Kloppenborg et al. 2008, Swan et al. 1998). Namely, midlife exposure to CVRs, including obesity, hypertension, and smoking, has been reported to markedly accelerate the deterioration of the executive functions and was believed to be a better predictor than exposure in later life (Tolppanen et al. 2012, Debette et al. 2011). Furthermore, a subtle decline in cognitive functions including cognitive reaction time and episodic memory, has been identified in middle-aged healthy subjects (younger than 50 years old), adding to the importance of midlife for the prevention of the pathological aspects of aging (Zimprich and Mascherek 2010, Ferreira et al. 2015). Findings as such have raised the question as to whether the observed declines in cognitive or executive abilities were mediated by the structural or functional alterations associated with aging or the exposure to other CVR factors in the cerebral vasculature during midlife. However, for the elucidation of mechanisms underlying those neurological disorders and the preservation of brain functions, a better and more comprehensive understanding of the structural modifications of the intracranial vasculature with aging is required. Nevertheless, previous studies investigating the effect of aging or general CVR factors on the intracranial vessel morphologies usually suffered from a relatively smaller sample size with a restricted age range, minimizing their power to detect earlier changes (Bullitt et al. 2010, Chen et al. 2019).

Therefore, in this study, we set out to address two questions. First, how does the characteristics of cerebral vasculature precipitated by MRA alter with aging. Thanks to improved MRA resolution and automatic vascular quantification, in vivo studies investigating the age-related morphological changes of the cerebral vasculature are possible. Second, whether aging affects the cerebral vascular structures differently in different age groups, especially in the middle-aged group (45-54 years), given changes in the cerebrovascular system during this period seemed to have marked predicting power for the cognitive outcomes in later life. In the current study, a large natural population in China aged from 35 to 75 was recruited. A more detailed division of subjects into subgroups based on age has been made to study the effect of aging as well as the conventional CVR factors on vascular morphologies in different age groups.

## Results

The participant characteristics are presented in Table 1. Figure 1 summarizes the recruitment in the current study. A total of 1240 participants were included; of these, 33 participants with poor image quality that is not adequate for vessel segmentation and 31 participants with incomplete cerebrovascular risk information were excluded. Therefore, 1176 participants were included in the subsequent analysis.

**Table 1.** Characteristics of the Study Participants.

| **Age Range** | 35-44 y | 45-54 y | 55-64 y | 65-75 y | Overall |
| --- | --- | --- | --- | --- | --- |
| **Demographic** |  |  |  |  |  |
| Participants, No. | 201 | 421 | 342 | 212 | 1176 |
| Age, mean (SD), years | 40.4 (3.3) | 51.1 (2.7) | 59.3 (2.8) | 70.3 (3.2) | 55.1 (9.9) |
| Female, No. (%) | 121 (0.60) | 279 (0.66) | 189 (0.55) | 113 (0.53) | 702 (0.60) |
| Body mass index, mean (SD) | 23.3 (3.0) | 23.5 (3.0) | 23.8 (2.9) | 23.2 (2.6) | 23.5 (2.9) |
| Smoking, No. (%) | 30 (0.15) | 87 (0.21) | 70 (0.20) | 39 (0.18) | 226 (0.19) |
| Alcohol consumption, No. (%) | 16 (0.08) | 42 (0.10) | 55 (0.16) | 24 (0.11) | 137 (0.12) |
| Diabetes diagnosis, No. (%) | 1 (0) | 29 (0.07) | 34 (0.10) | 30 (0.14) | 94 (0.08) |
| Hypertension, No. (%) | 22 (0.11) | 78 (0.19) | 117 (0.34) | 84 (0.40) | 301 (0.26) |
| Hyperlipidemia, No. (%) | 7 (0.03) | 14 (0.03) | 30.0 (0.09) | 15.0 (0.07) | 66 (0.06) |
| **Cerebrovascular morphology** |  |  |  |  |  |
| Vessel density, mean (SD) | 0.09 (0.02) | 0.08 (0.02) | 0.07 (0.02) | 0.07 (0.02) | 0.08 (0.02) |
| Vessel radius, mean (SD), mm | 0.78 (0.07) | 0.80 (0.08) | 0.82 (0.07) | 0.83 (0.07) | 0.81 (0.07) |
| Vessel tortuosity, mean (SD) | 1.19 (0.03) | 1.20 (0.03) | 1.20 (0.03) | 1.21 (0.03) | 1.2 (0.03) |

**Figure 1.**
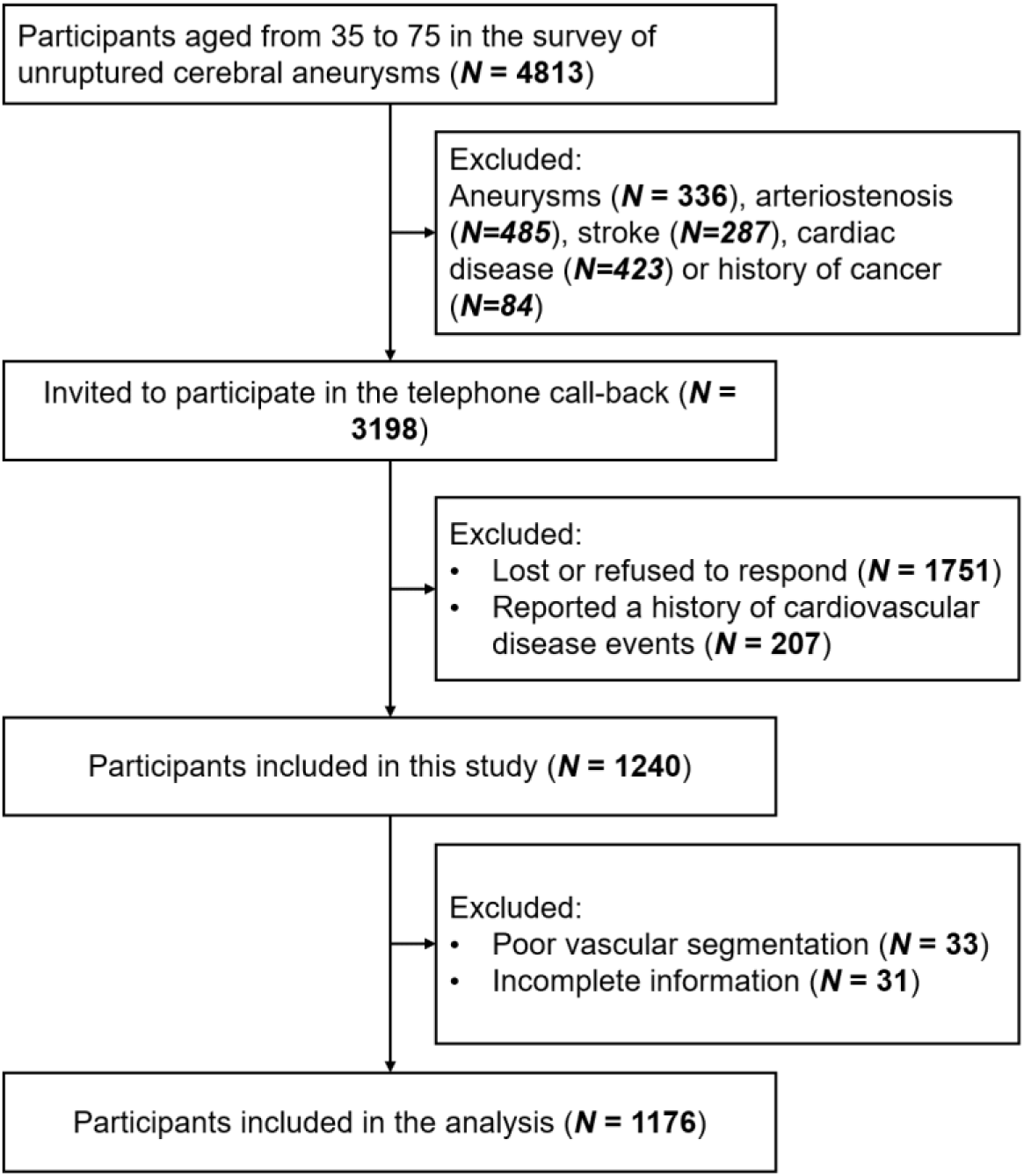
Flow chart of recruitment in the current study.

### Association between cerebrovascular morphology and aging

We first investigated the effect of age on the morphometric features of the cerebral vasculature in general. Age was found to be significantly associated with decreased vessel density (β=-5.7×10^−4^, 95% CI -0.001 to -4.3×10^−4^, p<0.001), increased vessel radius, (β=0.002, 95% CI 0.002 to 0.002, p<0.001), and increased vessel tortuosity, (β=8.5×10^−4^, 95% CI 6.7×10^−4^ to 0.001, p<0.001) after adjusting for CVR factors, as shown in Table 2. The participants were further divided into four groups based on their age to further examine whether the observed effect of aging differed in different age groups (Figure 2a-c). Surprisingly, vessel density was only found to be negatively associated with aging in participants aged 45-54, (β=-0.002, 95% CI -0.002 to -0.001, p<0.001), and 65-75, (β=-0.001, 95% CI -0.002 to -7.6×10^−5^, p=0.036), and only the association found in people aged 45-54 remained significant after adjusting for CVR factors, (β=-0.001, 95% CI -0.002 to -0.001, p=0.001), suggesting the effects of CVR factors are not negligible for people in the 65-75 age group, whereas the effect of aging dominates the changes of vascular features in the 45-54 age group. Similarly, increased vessel radius was only found to be associated with aging in participants aged 45-54, (β=0.005, 95% CI 0.002 to 0.008, p<0.001), and 65-75, (β=0.003, 95% CI 2.9×10^−4^ to 0.006, p=0.03), and, after adjusting for CVR factors, vessel radius was found to be significantly associated with advanced age only in participants aged 45-54, (β=0.005, 95% CI 0.003 to 0.008, p<0.001). Increased vessel tortuosity was found to be significantly associated with age in subjects aged 35-44 (β=0.001, 95% CI 0.001 to 0.003, p=0.03), and 45-54, (β=0.002, 95% CI 0.001 to 0.003, p=0.003). The associations remained significant after adjusting for CVR factors for participants aged 35-44, (β=0.002, 95% CI 3.4×10^−4^ to 0.003, p=0.01), and for participants aged 45-54, (β=0.001, 95% CI 4.4×10^−4^ to 0.003, p=0.006).

**Table 2.**
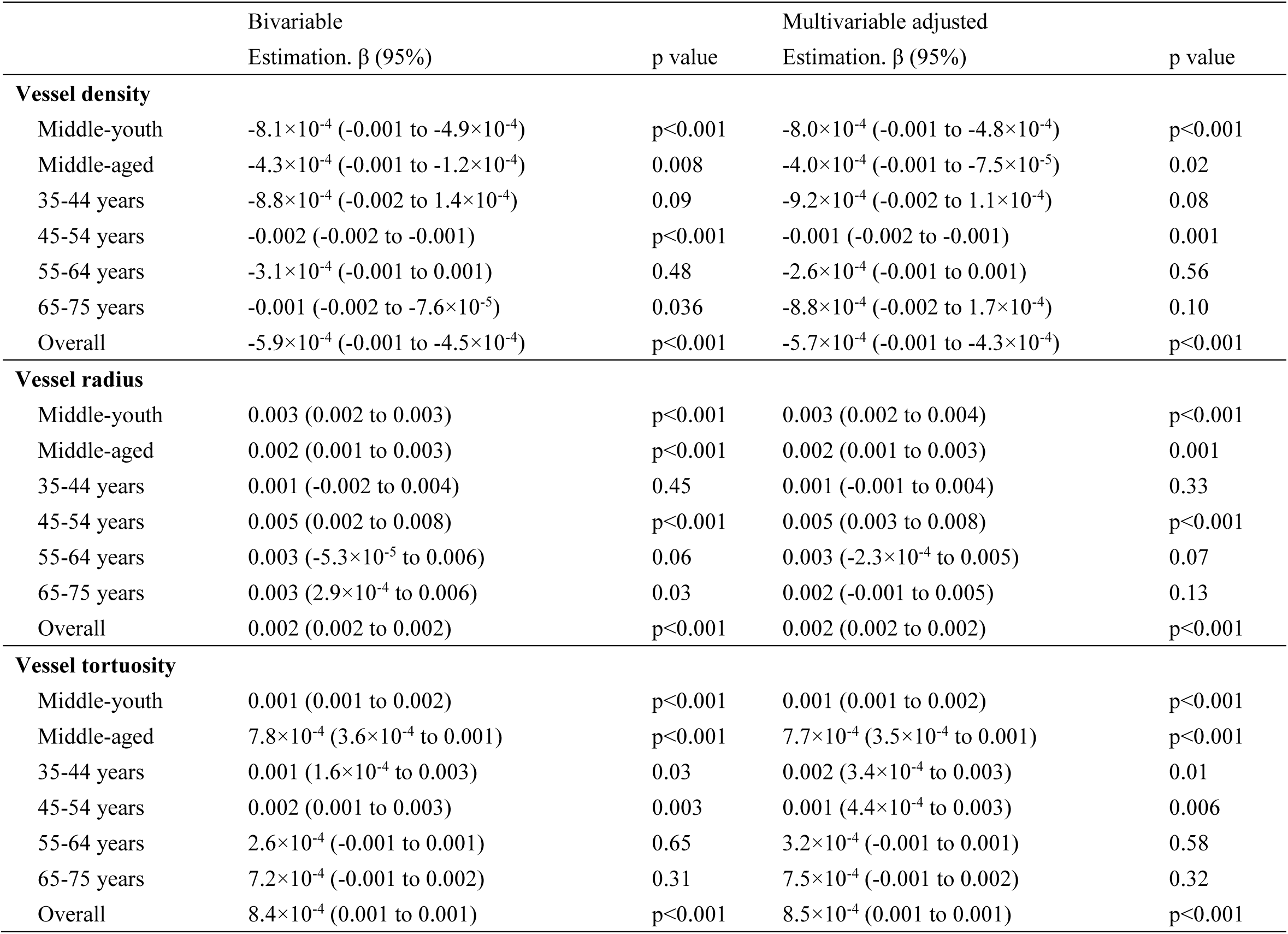
Association between age and cerebrovascular morphologies. In the multivariable adjusted models, sex and cerebrovascular risk factors were adjusted, including body mass index, smoking status, alcohol consumption, diabetes status, hypertension status and hyperlipidemia status. Middle-youth represented participants aged from 35 to 54 years. Middle-aged represented participants aged from 55 to 75 years.

|  | Bivariable |  | Multivariable adjusted |  |
| --- | --- | --- | --- | --- |
| | Estimation. $\beta$ (95%) | p value | Estimation. $\beta$ (95%) | p value |
| **Vessel density** |  |  |  |  |
| Middle-youth | $-8.1 \times 10^{-4}$ (-0.001 to $-4.9 \times 10^{-4}$) | $p < 0.001$ | $-8.0 \times 10^{-4}$ (-0.001 to $-4.8 \times 10^{-4}$) | $p < 0.001$ |
| Middle-aged | $-4.3 \times 10^{-4}$ (-0.001 to $-1.2 \times 10^{-4}$) | 0.008 | $-4.0 \times 10^{-4}$ (-0.001 to $-7.5 \times 10^{-5}$) | 0.02 |
| 35-44 years | $-8.8 \times 10^{-4}$ (-0.002 to $1.4 \times 10^{-4}$) | 0.09 | $-9.2 \times 10^{-4}$ (-0.002 to $1.1 \times 10^{-4}$) | 0.08 |
| 45-54 years | -0.002 (-0.002 to -0.001) | $p < 0.001$ | -0.001 (-0.002 to -0.001) | 0.001 |
| 55-64 years | $-3.1 \times 10^{-4}$ (-0.001 to 0.001) | 0.48 | $-2.6 \times 10^{-4}$ (-0.001 to 0.001) | 0.56 |
| 65-75 years | -0.001 (-0.002 to $-7.6 \times 10^{-5}$) | 0.036 | $-8.8 \times 10^{-4}$ (-0.002 to $1.7 \times 10^{-4}$) | 0.10 |
| Overall | $-5.9 \times 10^{-4}$ (-0.001 to $-4.5 \times 10^{-4}$) | $p < 0.001$ | $-5.7 \times 10^{-4}$ (-0.001 to $-4.3 \times 10^{-4}$) | $p < 0.001$ |
| **Vessel radius** |  |  |  |  |
| Middle-youth | 0.003 (0.002 to 0.003) | $p < 0.001$ | 0.003 (0.002 to 0.004) | $p < 0.001$ |
| Middle-aged | 0.002 (0.001 to 0.003) | $p < 0.001$ | 0.002 (0.001 to 0.003) | 0.001 |
| 35-44 years | 0.001 (-0.002 to 0.004) | 0.45 | 0.001 (-0.001 to 0.004) | 0.33 |
| 45-54 years | 0.005 (0.002 to 0.008) | $p < 0.001$ | 0.005 (0.003 to 0.008) | $p < 0.001$ |
| 55-64 years | 0.003 ($-5.3 \times 10^{-5}$ to 0.006) | 0.06 | 0.003 ($-2.3 \times 10^{-4}$ to 0.005) | 0.07 |
| 65-75 years | 0.003 ($2.9 \times 10^{-4}$ to 0.006) | 0.03 | 0.002 (-0.001 to 0.005) | 0.13 |
| Overall | 0.002 (0.002 to 0.002) | $p < 0.001$ | 0.002 (0.002 to 0.002) | $p < 0.001$ |
| **Vessel tortuosity** |  |  |  |  |
| Middle-youth | 0.001 (0.001 to 0.002) | $p < 0.001$ | 0.001 (0.001 to 0.002) | $p < 0.001$ |
| Middle-aged | $7.8 \times 10^{-4}$ ($3.6 \times 10^{-4}$ to 0.001) | $p < 0.001$ | $7.7 \times 10^{-4}$ ($3.5 \times 10^{-4}$ to 0.001) | $p < 0.001$ |
| 35-44 years | 0.001 ($1.6 \times 10^{-4}$ to 0.003) | 0.03 | 0.002 ($3.4 \times 10^{-4}$ to 0.003) | 0.01 |
| 45-54 years | 0.002 (0.001 to 0.003) | 0.003 | 0.001 ($4.4 \times 10^{-4}$ to 0.003) | 0.006 |
| 55-64 years | $2.6 \times 10^{-4}$ (-0.001 to 0.001) | 0.65 | $3.2 \times 10^{-4}$ (-0.001 to 0.001) | 0.58 |
| 65-75 years | $7.2 \times 10^{-4}$ (-0.001 to 0.002) | 0.31 | $7.5 \times 10^{-4}$ (-0.001 to 0.002) | 0.32 |
| Overall | $8.4 \times 10^{-4}$ (0.001 to 0.001) | $p < 0.001$ | $8.5 \times 10^{-4}$ (0.001 to 0.001) | $p < 0.001$ |

**Figure 2.**
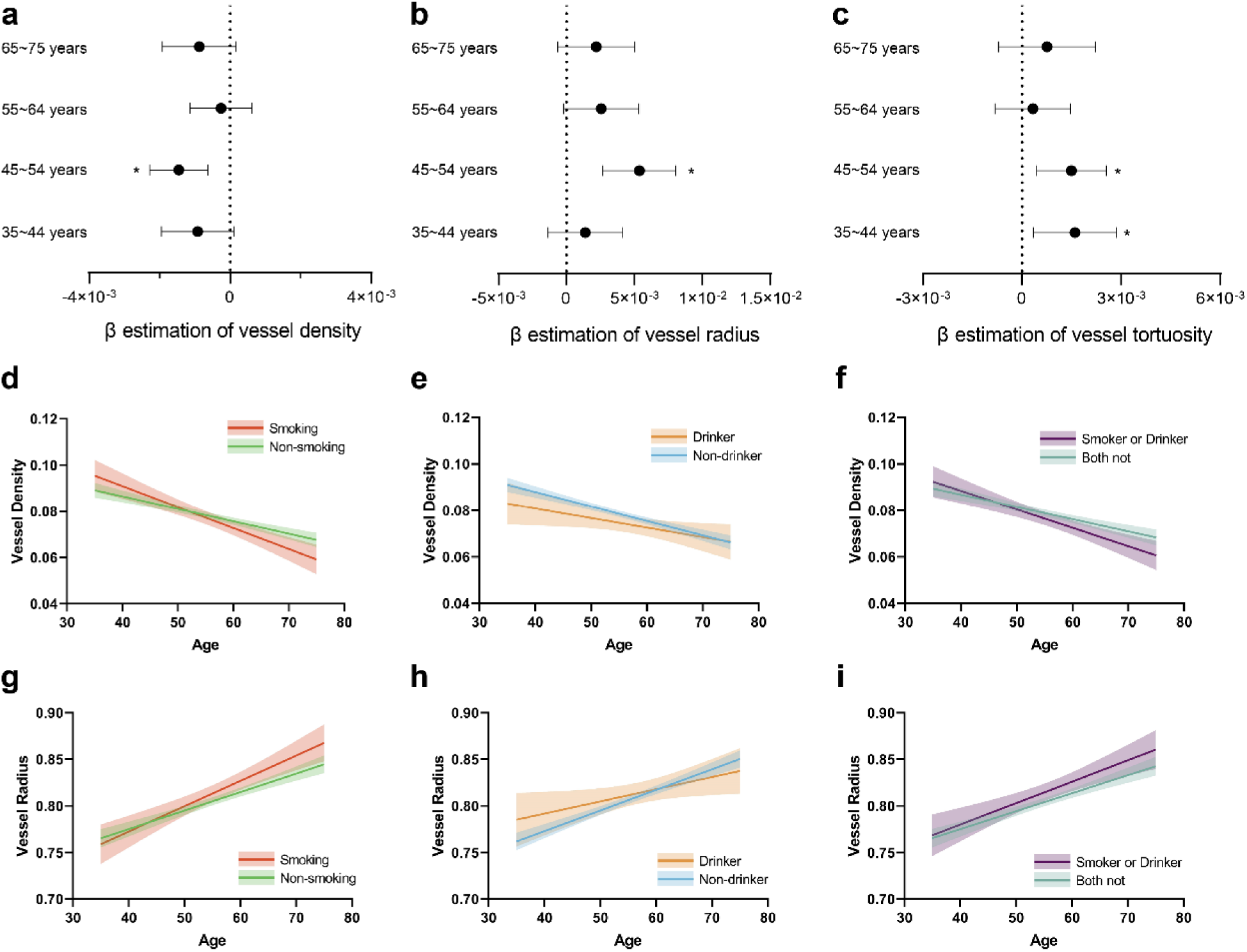
Association of cerebrovascular morphology with age and cerebrovascular risk factors. (a)∼(c) Regression results between age and cerebrovascular morphologies, including vessel density, vessel radius and vessel tortuosity, in different age groups. (d)∼(f) Association between vessel density and age in different groups (smoking and non-smoking, drinker and non-drinker, smoker or drinker and both not). Shaded area represents the 95% confidence interval. (g)∼(i) Association between vessel radius and age in different groups.

To better delineate the underlying mechanisms for dilated arteries, we further assessed the proportion of the smaller vessels visualized by MRA. As shown in Table 3, a general decrease in the proportion of the smaller vessels with advancing age was identified, (β= -3.7×10^−4^, 95% CI: -6.1×10^−4^, -1.3×10^−4^, p=0.03). The decrease in mean vessel radius reported before, however, was found not to be due to the decreased proportion of the smaller vessels, since the association between aging and vessel radius was not much affected after adjusting for the proportion of the smaller vessels, (β= 2.7×10^−4^, 95% CI: 6.6×10^−5^, 4.8×10^−4^, p=0.01), hence suggesting compensatory vasodilatation might be the cause here.

**Table 3.** Association between age and smaller vessels. In the multivariable adjusted models, sex and cerebrovascular risk factors were adjusted, including body mass index, smoking status, alcohol consumption, diabetes status, hypertension status and hyperlipidemia status.

| | Estimation, $\beta$ (95%CI) | p value |
| --- | --- | --- |
| **Proportion of smaller vessels (PSV)** |  |  |
| Bivariable | $-4.0 \times 10^{-4}$ ($-6.3 \times 10^{-4}$ to $-1.7 \times 10^{-4}$) | 0.001 |
| Multivariable adjusted | $-3.7 \times 10^{-4}$ ($-6.1 \times 10^{-4}$ to $-1.3 \times 10^{-4}$) | 0.003 |
| **Vessel Radius adjusted for PSV** | $2.7 \times 10^{-4}$ ($6.6 \times 10^{-5}$ to $4.8 \times 10^{-4}$) | 0.01 |

### Association between cerebrovascular morphologies and CVR factors

The relationships between cerebrovascular morphologies and CVR factors were further examined. Vessel radius and mean tortuosity were found not to be associated with any of the individual risk factors listed in Table 4. A decrease in vessel density was found in participants with diabetes after adjusting for age and other risk factors (β=-0.005, 95% CI -0.01 to -0.001, p<0.046). Interestingly, the interaction between age and smoking was found to significantly impact vessel density (β= -6.7×10^−4^, 95% CI: - 8.8×10^−4^, 4.6×10^−4^, p=0.002) and vessel radius (β= 0.002, 95% CI: 9.6×10^−4^, 0.002, p=0.01). However, the effect of the interaction between age and alcohol use seems to counteract the effect of aging alone on vascular density and vessel radius. While decreased vascular density and increased vessel radius were found in older adults in general, the interaction of age and alcohol consumption seems to result in a higher vascular density (β= 5.8×10^−4^, 95% CI: 8.6×10^−4^, 3.1×10^−4^, p=0.03) and a lower vessel radius (β= -0.002, 95% CI: -0.003, -8.6×10^−4^, p=0.04). In general, smoking was found to facilitate the deterioration of the vascular structures, while alcohol use seemed to decelerate the changes (Figure 2d-i). No significant interaction between age and other CVR factors was found.

**Table 4.** Association between cerebrovascular morphologies and cerebrovascular risk factors. Only significant interaction terms were presented. Middle-youth represented participants aged from 35 to 54 years. Middle-aged represented participants aged from 55 to 75 years.

|  | Bivariable |  | Multivariable |  |
| --- | --- | --- | --- | --- |
| | Estimation, $\beta$ (95%CI) | p value | Estimation, $\beta$ (95%CI) | p value |
| **Vessel density** |  |  |  |  |
| Age | $-5.9 \times 10^{-4}$ ($-7.2 \times 10^{-4}$ to $-4.5 \times 10^{-4}$) | $p < 0.001$ | $-5.7 \times 10^{-4}$ ($-7.1 \times 10^{-4}$ to $-4.3 \times 10^{-4}$) | $p < 0.001$ |
| Body mass index | $-2.4 \times 10^{-6}$ ($-4.8 \times 10^{-4}$ to $4.7 \times 10^{-4}$) | 0.99 | $1.4 \times 10^{-5}$ ($-4.6 \times 10^{-4}$ to $4.9 \times 10^{-4}$) | 0.95 |
| Smoking | -0.002 (-0.006 to 0.001) | 0.25 | $-2.7 \times 10^{-4}$ (-0.004 to 0.004) | 0.89 |
| Alcohol consumption | -0.004 (-0.009 to $-1.5 \times 10^{-4}$) | 0.043 | -0.003 (-0.008 to 0.001) | 0.16 |
| Diabetes diagnosis | -0.008 (-0.01 to -0.003) | 0.001 | -0.005 (-0.01 to $-9.1 \times 10^{-5}$) | 0.046 |
| Hypertension | -0.003 (-0.006 to $3.4 \times 10^{-4}$) | 0.08 | $5.8 \times 10^{-4}$ (-0.003 to 0.004) | 0.72 |
| Hyperlipidemia | 0.003 (-0.003 to 0.009) | 0.31 | 0.005 ($-5.4 \times 10^{-4}$ to 0.01) | 0.07 |
| Age*Smoking | -- | -- | $-6.7 \times 10^{-4}$ ($-8.8 \times 10^{-4}$ to $-4.6 \times 10^{-4}$) | 0.002 |
| Age*Alcohol consumption | -- | -- | $5.8 \times 10^{-4}$ ($8.6 \times 10^{-4}$ to $3.1 \times 10^{-4}$) | 0.03 |
| **Vessel radius** |  |  |  |  |
| Age | 0.002 (0.002 to 0.002) | $p < 0.001$ | 0.002 (0.002 to 0.002) | $p < 0.001$ |
| Body mass index | $3.6 \times 10^{-4}$ (-0.001 to 0.002) | 0.63 | $4.2 \times 10^{-4}$ (-0.001 to 0.002) | 0.56 |
| Smoking | 0.01 ($8.0 \times 10^{-4}$ to 0.02) | 0.035 | 0.01 (-0.001 to 0.02) | 0.08 |
| Alcohol consumption | 0.009 (-0.004 to 0.02) | 0.19 | $-7.7 \times 10^{-4}$ (-0.02 to 0.01) | 0.92 |
| Diabetes diagnosis | 0.02 (0.008 to 0.04) | 0.003 | 0.01 (-0.004 to 0.03) | 0.14 |
| Hypertension | 0.009 ($-3.5 \times 10^{-4}$ to 0.02) | 0.06 | -0.002 (-0.01 to 0.008) | 0.68 |
| Hyperlipidemia | -0.007 (-0.03 to 0.01) | 0.42 | -0.02 (-0.03 to 0.003) | 0.10 |
| Age*Smoking | -- | -- | 0.002 ($9.6 \times 10^{-4}$ to 0.002) | 0.01 |
| Age*Alcohol consumption | -- | -- | -0.002 (-0.003 to $-8.6 \times 10^{-4}$) | 0.036 |
| **Vessel tortuosity** |  |  |  |  |
| Age | $8.4 \times 10^{-4}$ ($6.7 \times 10^{-4}$ to 0.001) | $p < 0.001$ | $8.5 \times 10^{-4}$ ($6.7 \times 10^{-4}$ to 0.001) | $p < 0.001$ |
| Body mass index | $2.7 \times 10^{-4}$ ($-3.5 \times 10^{-4}$ to $8.8 \times 10^{-4}$) | 0.39 | $3.0 \times 10^{-4}$ ($-3.1 \times 10^{-4}$ to $9.1 \times 10^{-4}$) | 0.33 |
| Smoking | $9.3 \times 10^{-4}$ (-0.004 to 0.005) | 0.69 | 0.002 (-0.003 to 0.007) | 0.52 |
| Alcohol consumption | $-2.8 \times 10^{-4}$ (-0.006 to 0.005) | 0.92 | -0.003 (-0.009 to 0.004) | 0.40 |
| Diabetes diagnosis | 0.005 (-0.002 to 0.01) | 0.17 | $-6.2 \times 10^{-4}$ (-0.007 to 0.006) | 0.85 |
| Hypertension | 0.005 ($5.8 \times 10^{-4}$ to 0.009) | 0.03 | $-1.5 \times 10^{-4}$ (-0.004 to 0.004) | 0.95 |
| Hyperlipidemia | 0.002 (-0.005 to 0.01) | 0.57 | $-4.0 \times 10^{-4}$ (-0.008 to 0.007) | 0.92 |

### Association between cerebrovascular morphologies and CVR score

A cumulative CVR score of 0 to 6 was assigned accordingly, with a higher score indicating exposure to fewer CVR factors. The associations between CVR score and vascular morphologies have been reported in Table 5. Overall, a significant association was identified between CVR score and vessel radius after adjusting for age and sex (β= -0.004 95% CI: -0.008, -2.0×10^−4^, p=0.039). However, when participants were subdivided into age groups, CVR score was only found to be correlated with an increase in vascular density adjusted for age and sex (β= 0.002, 95% CI: 1.0×10^−4^, 0.004, p=0.041) in the 55-64 group, and a decrease in vessel radius after adjusting for age and sex (β= -0.010, 95% CI: -0.010, -1.6×10^−4^, p=0.048) in participants aged 35-44.

**Table 5.** Association between cerebrovascular score and cerebrovascular morphologies. Middle-youth represented participants aged from 35 to 54 years. Middle-aged represented participants aged from 55 to 75 years.

| | Estimation. $\beta$ (95%) | p value | Adjusted for age and sex<br>Estimation. $\beta$ (95%) | p value |
| --- | --- | --- | --- | --- |
| **Vessel density** |  |  |  |  |
| Middle-youth | $-1.9 \times 10^{-4}$ (-0.002 to 0.002) | 0.83 | $-6.3 \times 10^{-4}$ (-0.002 to 0.001) | 0.50 |
| Middle-aged | 0.002 (0.001 to 0.004) | 0.007 | 0.002 (0.001 to 0.004) | 0.01 |
| 35-44 years | 0.001 (-0.002 to 0.005) | 0.45 | 0.001 (-0.002 to 0.005) | 0.44 |
| 45-54 years | -0.001 (-0.004 to 0.001) | 0.18 | -0.001 (-0.003 to 0.001) | 0.28 |
| 55-64 years | 0.002 ($2.8 \times 10^{-4}$ to 0.004) | 0.03 | 0.002 ($1.0 \times 10^{-4}$ to 0.004) | 0.041 |
| 65-75 years | 0.003 ($-4.7 \times 10^{-4}$ to 0.006) | 0.10 | 0.002 (-0.001 to 0.005) | 0.17 |
| Overall | 0.002 (0.001 to 0.003) | 0.005 | $8.8 \times 10^{-4}$ ($-3.8 \times 10^{-4}$ to 0.002) | 0.17 |
| **Vessel radius** |  |  |  |  |
| Middle-youth | -0.004 (-0.010 to 0.001) | 0.14 | -0.003 (-0.009 to 0.002) | 0.22 |
| Middle-aged | -0.004 (-0.010 to 0.001) | 0.10 | -0.004 (-0.010 to 0.001) | 0.09 |
| 35-44 years | -0.009 (-0.02 to $5.4 \times 10^{-5}$) | 0.053 | -0.010 (-0.02 to $-1.6 \times 10^{-4}$) | 0.048 |
| 45-54 years | $-2.5 \times 10^{-4}$ (-0.007 to 0.007) | 0.94 | -0.001 (-0.008 to 0.005) | 0.69 |
| 55-64 years | -0.003 (-0.010 to 0.004) | 0.40 | -0.003 (-0.009 to 0.004) | 0.45 |
| 65-75 years | -0.008 (-0.02 to $2.8 \times 10^{-4}$) | 0.06 | -0.007 (-0.02 to 0.001) | 0.09 |
| Overall | -0.007 (-0.01 to -0.003) | 0.001 | -0.004 (-0.008 to $-2.0 \times 10^{-4}$) | 0.039 |
| **Vessel tortuosity** |  |  |  |  |
| Middle-youth | $-4.4 \times 10^{-4}$ (-0.003 to 0.002) | 0.71 | $5.1 \times 10^{-4}$ (-0.002 to 0.003) | 0.66 |
| Middle-aged | -0.001 (-0.003 to 0.001) | 0.38 | $-9.6 \times 10^{-4}$ (-0.003 to 0.001) | 0.42 |
| 35-44 years | -0.003 (-0.008 to 0.001) | 0.12 | -0.004 (-0.008 to $3.4 \times 10^{-4}$) | 0.07 |
| 45-54 years | 0.002 (-0.001 to 0.005) | 0.16 | 0.002 ($-3.6 \times 10^{-4}$ to 0.005) | 0.09 |
| 55-64 years | $-6.2 \times 10^{-4}$ (-0.003 to 0.002) | 0.65 | $-5.2 \times 10^{-4}$ (-0.003 to 0.002) | 0.71 |
| 65-75 years | -0.002 (-0.007 to 0.002) | 0.29 | -0.002 (-0.006 to 0.002) | 0.36 |
| Overall | -0.002 (-0.003 to $-1.4 \times 10^{-5}$) | 0.048 | $-2.6 \times 10^{-4}$ (-0.002 to 0.001) | 0.76 |

## Discussion

In this cross-sectional study, the effect of aging and other conventionally defined CVR factors on the intracerebral arterial morphologies acquired by 3D TOF MRA were investigated. As per previous studies, advanced age was found in our research to be associated with increased vessel radius, increased vessel tortuosity and decreased branch density (Bullitt et al. 2010, Chen et al. 2019). Thanks to the relatively large sample size of this study, assessments of the age- or CVR-related morphological alternations of the cerebral arteries through adulthood were carried out in four subgroups. Indeed, our study might be the first to report the changes in arterial morphologies in relation to aging in different age groups.

In subgroups, a significant age-related increase in tortuosity was first identified in the 35-44 age subgroup, appearing earlier than the age-related increase in mean arterial radius and decrease in branch density, both of which were first evident in the 45-54 age subgroup. In fact, aging was found to significantly influence all three vascular morphologies only in subjects aged 45-54, and such associations remained significant after adjusting for the vascular risk factors. On the contrary, although significant associations of aging with branch density and mean vascular radius were identified in the 65-75 subgroup initially, those ceased to be significant after adjusting for the vascular risk factors. Our results combined have at least two implications. First, tortuosity may be a more sensitive metrics for vascular aging since it appears to be changing rapidly with aging, even in the youngest subjects. Second, while aging dominates the changes of the morphological features reported above in the 45-54 subgroup, common vascular risk factors may have played a more crucial part in the 65-75 subgroup. However, since a full history of chronic conditions from the patients was unavailable, and the vascular risk factors were dichotomously coded (1 for the exposure of risk and 0 for the absence), the investigation of the effect of the chronic conditions duration on vessel structures was not feasible. Further studies may benefit from having such information. For example, it is possible that the effects of the common vascular risks, gradually re-shaping the cerebral vascular system, accrue through a lifetime until significant changes can be observed.

Overall, the observed age-related morphological alternations may reflect impaired vascular functions. First, since tortuosity increases the total vessel length, leading to increased flow resistance or, in advanced cases, occlusion of blood flow, it may underpin the previously reported age-related reduction in cerebral perfusion (Han 2012, Melamed et al. 1980). Indeed, tortuosity has been considered to increase the risk of stroke (Alazzaz et al. 2000, Diedrich et al. 2011). Moreover, a recent study found that the tortuosity index of the intracranial vessels can be used to differentiate healthy individuals from subjects with mild cognitive impairment or AD, suggesting that tortuosity may also reflect some pathological changes in the brain (Barbara-Morales et al. 2020). Second, due to the failure of the current MRA to detect under-perfused vessels, the age-related reduction in the branch density may also be the result of impaired cerebral perfusion. Nevertheless, as suggested previously (Bullitt et al. 2010), such reduction in branch density with age may also result from a physical loss of smaller vessels. In view of the restrictions posed by the current imaging method, it is not plausible to attribute the observed reduction in branch density to either one of the causes. Nonetheless, the fact that lower branch density was identified in the older population suggests decrements in vascular functions in the aging population. Third, our results also showed that advanced age was significantly associated with an increase in the mean vessel radius, in agreement with the results from previous MRA studies (Bullitt et al. 2010, Chen et al. 2019). Such results may arise from the rarefaction of smaller branches, resulting in a relative increase in the proportion of larger branches, compensatory vasodilation, or both. Our results, however, suggest that compensatory vessel dilation may be a more viable explanation, since aging was still significantly associated with increased vessel density after adjusting for the proportion of smaller vessels. As mentioned before, tortuous vessels increase the flow resistance, resulting in a more prominent drop in the affected sections of vessels and eventually leading to decreased cerebral flow in the downstream vessels. Although very tempting, age-related compensatory vasodilation, which, as indicated by our results, appears later than age-related alternation in the mean vessel tortuosity, may work to counteract the pressure drop brought about by increased tortuosity, and thus maintaining a constant blood flow in the downstream parts. It is even possible that early intervention methods that either prevent or slow further increase of tortuosity may be effective for the preservation of normal vascular functions. Taken together, our results may imply an age-related disturbance of the cerebral hemostasis, which may underlie age-related brain pathologies and would have important implications for early clinical prevention or intervention methods targeting the middle-aged population.

In the current study, vascular risk factors were less related to cerebrovascular morphologies after adjusting for age. Whereas having less vascular risks (indicated by a higher CVR score) might mitigate the aging-related morphological alterations, as we found that vascular score was positively correlated with vascular density at the age of 55-75. Our results combined emphasized that the common vascular risk factors may play a more prominent role in aged groups. The research in young adults has shown similar findings that lower modifiable cardiovascular score (more cardiovascular risk factors) is associated with lower cerebral vascular density, raising the potential that some individuals may be starting to diverge to different risk trajectories for brain vascular health in early adulthood(Williamson et al. 2018). These results further underline the importance of managing CVR to protect the health of cerebrovascular system. Since reducing vascular risk factors and sustained lifestyle intervention can reduce cardiovascular burden (Spring et al. 2014, Shah et al. 2016), and improve cerebral blood flow (Espeland et al. 2018).

Interaction of aging and risk factors on vascular morphology was presented. Greater effects of aging on cerebrovascular morphologies in smokers were observed in this study. A previous report has indicated that cigarette smoking could induce vascular smooth muscle cell senescence (Centner, Bhide and Salazar 2020). The accumulation of senescent VSMCs with aging may be the underlying cause for our observation. The interaction of aging with alcohol consumption also significantly affects the vascular morphologies was the. Although the relationship between aging and vascular morphology was steeper in non-drinking participants, vascular density was lower in drinking participants for almost the entire age range. The possible explanation is that few participants are drinkers in our study, leading to the minimal power of our results. Future studies with more participants would help verify these interactions.

This study suffered from several limitations: 1) only one MRI modality, TOF MRA, was available in the study, which makes further investigation of the potential causes of the discussed vascular alterations implausible. For example, to extract the general trend of the age-related cerebral perfusion changes in different age subgroups, arterial spin-labeling would be necessary. 2) Risk factors were coded in our analysis as only having dichotomous values indicating whether the subject had the chronic condition at the time when their data was collected, and their medical history was not available. However, as implied by our results, the effect of those risk factors may accumulate throughout time until the consequence is noticeable. Future studies would benefit from having access to the medical history and more detailed information of the subjects’ chronic conditions such as the duration. 3) The cerebral vessel structural information obtained from TOF MRA was averaged for each individual, and thus might be unable to identify subtle correlations between aging and local vessels. Existing studies have also primarily focused on the cerebral vasculature in terms of whole-brain averaging or local averaging (Williamson et al. 2018, Mouches et al. 2021), since it is laborious to distinguish vessels by anatomical structure on images manually. More intelligent techniques, such as machine learning-based approaches, are needed to automate the distinction between different cerebral vessels.

In the current study, we present an analysis of the 3D TOF MRA images of 1176 subjects recruited from 2 communities in Shanghai. Significant differences in the cerebrovascular morphologies were identified between older subjects and younger subjects even after adjusting for vascular risk factors. In subgroups based on age, age was a significant predictor for the alternations of all three vessel morphologies extracted by MRA only in the 45-54 subgroup. In the 65-75 subgroup, although significant associations of aging with branch density and vessel radius were initially identified, the significance has ceased to exist after adjusting for vascular risk factors. Furthermore, the age-related increase in tortuosity was the only significant age-related change identified in the youngest subgroup. These results provide two key insights. First, morphometric changes visualizable by MRA may underlie different physiological processes. Second, early intervention methods that either prevent or slow further increases of tortuosity may be effective for preserving normal vascular functions.

## Materials and Methods

### Participants

Participants in the present study also completed the survey of unruptured cerebral aneurysms in Chinese adults (Li et al. 2013). This cross-sectional study initially enrolled 4813 participants (aged 35 to 75 years) who completed the survey between June 2007 and June 2011 from two different Shanghai communities in China. Ten years later (2017 to 2021), participants were invited to call back and reported on their health status. As shown in Figure 1, 3198 participants at the first survey who were free of cardiac disease, stroke, arteriostenosis or a history of cancer were first selected for the call back. Participants with identified intracranial aneurysms were not included to prevent potential influence of cerebral aneurysms on vascular morphology. Among the 3198 subjects, 1751 participants that either withdrew or refused to respond to the call back and 207 participants who had severe cardiovascular disease (CVD) events, including myocardial infarction, unstable angina, heart failure, coronary heart disease or stroke over the 10 years were excluded. 1240 participants were included in final the analysis. In these participants, although some have suffered chronic conditions, such as hypertension and diabetes, these conditions were well managed, and no severe CVD events were reported during the past ten years. The study was approved by the institutional review board of the Sixth People’s Hospital, Shanghai, China. All participants were voluntarily enrolled and provided written informed consent before data collection.

### Assessment of Risk Factors

Initially, all participants were asked to complete a standard questionnaire to provide demographic characteristics, personal and family medical history, and lifestyle risk factors. Information about smoking, alcohol consumption, hypertension, diabetes, hyperlipidemia, stroke, coronary heart disease, myocardial infarction, and arrhythmia was obtained. In addition, participants completed physical examinations including measurement of height, weight, and blood pressure. Venous blood samples were collected by certified nurses between 7 AM and 8 AM after overnight fasting. The presence of hypertension was defined as systolic blood pressure ≥ 140 mm Hg or diastolic blood pressure ≥ 90 mm Hg, a previous diagnosis of hypertension, or use of anti-hypertensive medications. Smokers were defined as smoked>100 cigarettes in a lifetime, and drinker was defined as consuming >30 g of alcohol per week for 1 or more year. The presence of diabetes was defined by a fasting level of plasma glucose ≥7.0 mmol/L, a previous diagnosis of diabetes, or treatment with antidiabetic drugs. Hyperlipidemia was defined by total plasma cholesterol (TC) ≥ 5.2 mmol/L, triglycerides (TG) ≥ 1.7 mmol/L, a previous diagnosis of hyperlipidemia, or use of lipid-lowering drugs. During the call back, a new questionnaire was administered by trained staff to obtain information on medical history within the past 10 years. Due to the limitations of the call back, accurate assessments of the vascular risk factors were not possible. Hence the vascular risk information from baseline was used in the subsequent analysis.

### MRI Scanning and Processing

All MRI examinations were performed on one 3.0T MRI system (Philips Healthcare, Amsterdam, The Netherlands). 3D-TOF MRA was completed using 3-dimensional T1-weighted fast-field sequences with the following settings: repetition time/echo time, 35/7; flip angle, 20 degrees; field of view, 250×190×108 mm3; 4 slabs (180 slices); slice thickness, 0.8 mm; matrix, 732×1024; acquisition time, 8 minutes and 56 seconds. Any adverse events during the MRI examination, including panic attack, claustrophobia, dizziness, or falling, were queried and monitored by the investigators. Cerebral vessel segmentation and morphologies extraction were completed on TOF MRA with the following steps: 1) vascular structures were enhanced using the Jerman Enhancement Filter (Jerman et al. 2016); 2) the level-set based method was used to obtain vessel segmentation; 3) segmentation results were firstly checked by B.Y.Z and B.W. Segmentations with too few vascular branches, nonvascular tissue remnants, or incorrect vascular branch adhesions were re-segmented by adjusting the algorithm parameters. Then all segmentation results were visually checked twice by two radiologist J.L and Y.H.L who have over 3 years’ experience with neuroimaging interpretation. The segmented vessels that were still showing too few branches, remnants of nonvascular tissue, or severe vascular branch adhesions were defined as poor vascular segmentation and excluded from the analysis; 4) vessel centerlines were extracted using the Skeleton 3D toolbox(Kollmannsberger et al. 2017); 5) cerebrovascular morphologic measurements, including vessel density, radius and tortuosity were calculated. To avoid the influence of extracerebral vessels, the brain extraction tool of FSL (https://fsl.fmrib.ox.ac.uk/fsl/fslwiki/) was used to segment the brain regions on MRA images. Only vessels involved in the brain region were included in the subsequent analysis. The vessel density was defined as the ratio of intracranial artery branch numbers to the brain volume. The vessel tortuosity represents the averaged tortuosity for each segment and was calculated as the ratio of the shortest path and the actual length of a given vessel segment. The vessel radius was determined in each centerline points and then averaged over each individual participant. The smaller vessels were defined as vessels with a mean radius less than a certain threshold (global thresholding based on the radius distribution of the whole intracranial arteries) (Zhang et al. 2021). All data processing processes were accomplished on the Matlab2019a (MathWorks, Natick, Massachusetts, USA).

### Statistical Analysis

Bivariable and multivariable analyses were completed to investigate association between cerebrovascular morphologies (cerebral vessel density, radius and tortuosity) and vascular risk makers (age, body mass index, smoking, alcohol consumption, diabetes, hypertension and hyperlipidemia). Linear regression models were used with the risk makers as the determinant and cerebrovascular morphologies as outcome variables. The differential influence of aging on cerebrovascular morphologies by other risk makers was tested using interaction terms in the multivariable model. Participants were divided into different age groups according to their age: middle-youth (35-54 years), middle-aged (55-75 years), 35 to 44 years, 45 to 54 years, 55 to 64 years and 65 to 75 years.

The individuals’ combined CVR score from 6 risk factors was also analyzed, as existing evidence indicated combined risk factor score was correlated with the brain health in young adults (Williamson et al. 2018). The score was cumulative based on each of the following factors: body mass index ≤ 24; no smoking; not a drinker; no hyperlipidemia; no hypertension; no diabetes, with a higher number indicating less exposure to cerebrovascular risks. The relationships between the CVR score and MRA findings were studied using the linear regression model. In addition, linear analysis was completed to investigate the correlations between aging and cerebrovascular morphologies in different age subgroups.

All statistical analysis was completed using Matlab2019a (MathWorks, Natick, Massachusetts, USA). Shapiro-Wilk test was performed to confirm the normality of continuous variables. Results of normally distributed continuous variables are presented as mean and standard deviation (SD) or as median and interquartile range (IQR). For categorical variables, frequencies, and proportions are presented. The two-sample t-test was used to evaluate group differences. All tests were 2-sided and a probability value <0.05 was considered significant.

## Data Availability

All data are available on reasonable request to corresponding authors.

## Reference

Alazzaz, A., J. Thornton, V. A. Aletich, G. M. Debrun, J. I. Ausman & F. Charbel (2000) Intracranial Percutaneous Transluminal Angioplasty for Arteriosclerotic Stenosis. Archives of Neurology, 57, 1625–1630.

Alber, J., S. Alladi, H.-J. Bae, D. A. Barton, L. A. Beckett, J. M. Bell, S. E. Berman, G. J. Biessels, S. E. Black, I. Bos, G. L. Bowman, E. Brai, A. M. Brickman, B. L. Callahan, R. A. Corriveau, S. Fossati, R. F. Gottesman, D. R. Gustafson, V. Hachinski, K. M. Hayden, A. M. Helman, T. M. Hughes, J. D. Isaacs, A. L. Jefferson, S. C. Johnson, A. Kapasi, S. Kern, J. C. Kwon, J. Kukolja, A. Lee, S. N. Lockhart, A. Murray, K. E. Osborn, M. C. Power, B. R. Price, H. F. M. Rhodius-Meester, J. A. Rondeau, A. C. Rosen, D. L. Rosene, J. A. Schneider, H. Scholtzova, C. E. Shaaban, N. C. B. S. Silva, H. M. Snyder, W. Swardfager, A. M. Troen, S. J. van Veluw, P. Vemuri, A. Wallin, C. Wellington, D. M. Wilcock, S. X. Xie & A. H. Hainsworth (2019) White matter hyperintensities in vascular contributions to cognitive impairment and dementia (VCID): Knowledge gaps and opportunities. Alzheimer’s & dementia (New York, N. Y.), 5, 107–117.

Banerjee, G., D. Wilson, H. R. Jäger & D. J. Werring (2016) Novel imaging techniques in cerebral small vessel diseases and vascular cognitive impairment. Biochim Biophys Acta, 1862, 926–38.

Barbara-Morales, E., J. Perez-Gonzalez, K. C. Rojas-Saavedra & V. Medina-Banuelos (2020) Evaluation of Brain Tortuosity Measurement for the Automatic Multimodal Classification of Subjects with Alzheimer’s Disease. Computational Intelligence and Neuroscience, 2020.

Bullitt, E., D. Zeng, B. Mortamet, A. Ghosh, S. R. Aylward, W. Lin, B. L. Marks & K. Smith (2010) The effects of healthy aging on intracerebral blood vessels visualized by magnetic resonance angiography. Neurobiology of Aging, 31, 290–300.

Centner, A. M., P. G. Bhide & G. Salazar (2020) Nicotine in Senescence and Atherosclerosis. Cells, 9.

Chen, L., J. Sun, D. S. Hippe, N. Balu, Q. Yuan, I. Yuan, X. Zhao, R. Li, L. He, T. S. Hatsukami, J.-N. Hwang & C. Yuan (2019) Quantitative assessment of the intracranial vasculature in an older adult population using iCafe. Neurobiology of Aging, 79, 59–65.

D’Agostino, R. B., R. S. Vasan, M. J. Pencina, P. A. Wolf, M. Cobain, J. M. Massaro & W. B. Kannel (2008) General Cardiovascular Risk Profile for Use in Primary Care. Circulation, 117, 743–753.

Debette, S., S. Seshadri, A. Beiser, R. Au, J. J. Himali, C. Palumbo, P. A. Wolf & C. DeCarli (2011) Midlife vascular risk factor exposure accelerates structural brain aging and cognitive decline. Neurology, 77, 461–468.

Diedrich, K. T., J. A. Roberts, R. H. Schmidt, C. K. Kang, Z. H. Cho & D. L. Parker (2011) Validation of an arterial tortuosity measure with application to hypertension collection of clinical hypertensive patients. BMC Bioinformatics, 12 Suppl 10, S15.

Donato, A. J., D. R. Machin & L. A. Lesniewski (2018) Mechanisms of Dysfunction in the Aging Vasculature and Role in Age-Related Disease. Circulation Research, 123, 825–848.

Espeland, M. A., J. A. Luchsinger, R. H. Neiberg, O. Carmichael, P. J. Laurienti, X. Pi-Sunyer, R. R. Wing, D. Cook, E. Horton, R. Casanova, K. Erickson, R. N. Bryan & M. Action Hlth Diabet Brain (2018) Long Term Effect of Intensive Lifestyle Intervention on Cerebral Blood Flow. Journal of the American Geriatrics Society, 66, 120–126.

Ferreira, D., R. Correia, A. Nieto, A. Machado, Y. Molina & J. Barroso (2015) Cognitive decline before the age of 50 can be detected with sensitive cognitive measures. Psicothema, 27, 216–22.

Han, H.-C. (2012) Twisted blood vessels: symptoms, etiology and biomechanical mechanisms. Journal of vascular research, 49, 185–197.

Jerman, T., F. Pernus, B. Likar & Z. Spiclin (2016) Enhancement of Vascular Structures in 3D and 2D Angiographic Images. Ieee Transactions on Medical Imaging, 35, 2107–2118.

Kalaria, R. N. (2002) Small Vessel Disease and Alzheimer’s Dementia: Pathological Considerations. Cerebrovascular Diseases, 13(suppl 2), 48–52.

Kloppenborg, R. P., E. van den Berg, L. J. Kappelle & G. J. Biessels (2008) Diabetes and other vascular risk factors for dementia: which factor matters most? A systematic review. Eur J Pharmacol, 585, 97–108.

Kollmannsberger, P., M. Kerschnitzki, F. Repp, W. Wagermaier, R. Weinkamer & P. Fratzl (2017) The small world of osteocytes: connectomics of the lacuno-canalicular network in bone. New Journal of Physics, 19.

Laurent, S. & P. Boutouyrie (2015) The Structural Factor of Hypertension Large and Small Artery Alterations. Circulation Research, 116, 1007–1021.

Leeuwis, A. E., M. R. Benedictus, J. P. A. Kuijer, M. A. A. Binnewijzend, A. M. Hooghiemstra, S. C. J. Verfaillie, T. Koene, P. Scheltens, F. Barkhof, N. D. Prins & W. M. van der Flier (2017) Lower cerebral blood flow is associated with impairment in multiple cognitive domains in Alzheimer’s disease. Alzheimers & Dementia, 13, 531–540.

Li, M.-H., S.-W. Chen, Y.-D. Li, Y.-C. Chen, Y.-S. Cheng, D.-J. Hu, H.-Q. Tan, Q. Wu, W. Wang, Z.-K. Sun, X.-E. Wei, J.-Y. Zhang, R.-H. Qiao, W.-H. Zong, Y. Zhang, W. Lou, Z.-Y. Chen, Y. Zhu, D.-R. Peng, S.-X. Ding, X.-F. Xu, X.-H. Hou & W.-P. Jia (2013) Prevalence of Unruptured Cerebral Aneurysms in Chinese Adults Aged 35 to 75 Years. Annals of Internal Medicine, 159, 514-+.

Melamed, E., S. Lavy, S. Bentin, G. Cooper & Y. Rinot (1980) REDUCTION IN REGIONAL CEREBRAL BLOOD-FLOW DURING NORMAL AGING IN MAN. Stroke, 11, 31–35.

Mouches, P., S. Langner, M. Domin, M. D. Hill & N. D. Forkert (2021) Influence of cardiovascular riskfactors on morphological changes of cerebral arteries in healthy adults across the life span. Scientific Reports, 11.

Shah, R. V., V. L. Murthy, L. A. Colangelo, J. Reis, B. A. Venkatesh, R. Sharma, S. A. Abbasi, D. C. Goff, J. J. Carr, J. S. Rana, J. G. Terry, C. Bouchard, M. A. Sarzynski, A. Eisman, T. Neilan, S. Das, M. Jerosch-Herold, C. E. Lewis, M. Carnethon, G. D. Lewis & J. A. C. Lima (2016) Association of Fitness in Young Adulthood With Survival and Cardiovascular Risk The Coronary Artery Risk Development in Young Adults (CARDIA) Study. Jama Internal Medicine, 176, 87–95.

Silvestrini, M., P. Pasqualetti, R. Baruffaldi, M. Bartolini, Y. Handouk, M. Matteis, F. Moffa, L. Provinciali & F. Vernieri (2006) Cerebrovascular Reactivity and Cognitive Decline in Patients With Alzheimer Disease. Stroke, 37, 1010–1015.

Spring, B., A. C. Moller, L. A. Colangelo, J. Siddique, M. Roehrig, M. L. Daviglus, J. F. Polak, J. P. Reis, S. Sidney & K. Liu (2014) Healthy Lifestyle Change and Subclinical Atherosclerosis in Young Adults Coronary Artery Risk Development in Young Adults (CARDIA) Study. Circulation, 130, 10-+.

Swan, G. E., C. DeCarli, B. L. Miller, T. Reed, P. A. Wolf, L. M. Jack & D. Carmelli (1998) Association of midlife blood pressure to late-life cognitive decline and brain morphology. Neurology, 51, 986–993.

Tolppanen, A. M., A. Solomon, H. Soininen & M. Kivipelto (2012) Midlife vascular risk factors and Alzheimer’s disease: evidence from epidemiological studies. J Alzheimers Dis, 32, 531–40.

Walker, K. A., A. R. Sharrett, A. Wu, A. L. C. Schneider, M. Albert, P. L. Lutsey, K. Bandeen-Roche, J. Coresh, A. L. Gross, B. G. Windham, D. S. Knopman, M. C. Power, A. M. Rawlings, T. H. Mosley & R. F. Gottesman (2019) Association of Midlife to Late-Life Blood Pressure Patterns With Incident Dementia. Jama-Journal of the American Medical Association, 322, 535–545.

Williamson, W., A. J. Lewandowski, N. D. Forkert, L. Griffanti, T. W. Okell, J. Betts, H. Boardman, T. Siepmann, D. McKean, O. Huckstep, J. M. Francis, S. Neubauer, R. Phellan, M. Jenkinson, A. Doherty, H. Dawes, E. Frangou, C. Malamateniou, C. Foster & P. Leeson (2018) Association of Cardiovascular Risk Factors With MRI Indices of Cerebrovascular Structure and Function and White Matter Hyperintensities in Young Adults. Jama-Journal of the American Medical Association, 320, 665–673.

Xu, X., B. Wang, C. Ren, J. Hu, D. A. Greenberg, T. Chen, L. Xie & K. Jin (2017) Age-related Impairment of Vascular Structure and Functions. Aging and Disease, 8, 590–610.

Zhang, B., Y. Wang, B. Wang, Y.-H. Chu, Y. Jiang, M. Cui, H. Wang & X. Chen (2021) MRI-Based Investigation of Association Between Cerebrovascular Structural Alteration and White Matter Hyperintensity Induced by High Blood Pressure. Journal of Magnetic Resonance Imaging.

Zhang, D. & M. E. Raichle (2010) Disease and the brain’s dark energy. Nature Reviews Neurology, 6, 15–28.

Zimprich, D. & A. Mascherek (2010) Five views of a secret: does cognition change during middle adulthood? European Journal of Ageing, 7, 135–146.

